# VAGINAL STEAMING PRACTICES AMONG WOMEN AT MULAGO STI CLINIC, UGANDA: PREVALENCE, LACTOBACILLI PROPORTION DIFFERENCES, AND ASSOCIATED FACTORS

**DOI:** 10.64898/2026.08.28.26361407

**Authors:** Dorothy Kakai, Twinamasiko Nelson, Namutale Racheal, Kigozi Enos, Akinyi Lilian, Bagaya Jamidah, Mutesi Bushira, Kajumbula Henry, Sarah Nakubulwa

## Abstract

**Background:** Vaginal steaming has gained popularity among women for reasons known best to them. However, the effects of vaginal steaming on vaginal lactobacilli levels remain poorly understood and understudied. This study investigated the prevalence, assessed differences in the presence of bacterial vaginosis (BV) among women who practiced vaginal steaming and those who do not and determined the factors associated with vaginal steaming among women attending Mulago Sexually Transmitted Infections’ (STI) clinic in Uganda.

**Methods:** This study utilized a cross-sectional design to enroll 181 women aged 18 to 49 years who were systematically sampled at Mulago STI clinic. Interviews were conducted to obtain the demographic characteristics of the participants. Vaginal swabbing and Gram staining were done to attain lactobacilli counts by microscopy which were categorized using the Nugent score. Data were analyzed using Stata. The potential confounding effects of other variables on the relation between vaginal steaming and the presence of bacterial vaginosis as well as factors associated with vaginal steaming were assessed using modified Poisson regression.

**Results:** Prevalence of vaginal steaming was 40.3%, (95% confidence interval (CI) 33.0% - 48.0%). There were 41.1% women who practiced vaginal steaming occasionally, 53.4% who used hot water having herbs and 78.1% who practiced vaginal steaming for medical reasons. There was no difference in the presence of bacterial vaginosis when women who practiced vaginal steaming were compared to those who did not (p-value = 0.286). Factors that were significantly associated with vaginal steaming included having experienced vaginal issues (aPR = 0.07, 95% CI 0.01 - 0.12, p value = < 0.001) and contraceptive use (aPR= 0.52, 95% CI 0.37 - 0.72, p value = 0.001).

**Conclusions:** About 2 in every 5 women at Mulago STI clinic reported to have indulged in vaginal steaming. There was no difference in the presence of bacterial vaginosis when women who practiced vaginal steaming were compared to those who did not. Having experienced vaginal issues and contraceptive use were significantly associated with vaginal steaming among women at Mulago STI clinic, Uganda.

The Ministry of Health of Uganda should establish targeted screening and treatment for bacterial vaginosis alongside other sexually transmitted infections.

## Background

Vaginal steaming is a practice that involves exposing the female genital area to steam from either plain hot water or hot water infused with various herb(s). The practice has become popular globally among women as it is done for hygiene purposes considered as being feminine [1]. However, the burden of vaginal steaming has not been reported globally. The prevalence of intra-vaginal practices among women in Chonburi is 10.9% [2]. More than 95% of Malawian Women use at least one intra vaginal practice (Esber, Rao et al. 2016). In southwestern Uganda, 47.2% of the women use intravaginal practices [3]. There are several factors that are associated with vaginal steaming including age, sex, education, ethnicity among others.

A study done among Suriname women showed that education level was associated with vaginal herb use [4]. A 2015 multi-country systematic review done in Australia, Canada, New Zealand, the United Kingdom, the United States of America revealed that vaginal steaming practices are done for hygiene purposes and the practice is deeply rooted in cultural traditions [5]. Grey literature shows that some women practice vaginal steaming for medical reasons [6]. The vaginal microbiota is dominated by lactobacilli [7] since the vaginal pH is naturally acidic [8]. Vaginal steaming may alter the vaginal microbiota and increase the risk of vaginal infections [9]. The change in vaginal microbiota and pH causes loss of many *Lactobacillus* species which alters the vaginal ecosystem hence causing an alteration in the vaginal lactobacilli count [10].

Vaginal *Lactobacillus* is a genus of gram-positive, rod-shaped, non-spore-forming bacteria that predominates the vaginal microbiome [11]. Lactobacilli count of women aged 18-49 are affected by sexually transmitted infections (*Neisseria gonorrhoeae, Chlamydia trachomatis*, and *Trichomonas vaginalis*) and some antibiotics [12–14]. Research shows that the possible cause of bacterial vaginosis (BV) are those factors which cause the vaginal pH to increase thereby leading to a decrease in lactobacilli and an increase in concentration of anaerobic bacteria hence cascade of population changes that imply bacterial vaginosis [15].

In 2023, World Health Organization reported incidences of over1 million sexually transmitted infections globally among adults aged 15-49 annually: 129 million cases of chlamydia, 82 million cases of gonorrhea, 7.1 million cases of syphilis [16]. Globally, there is a 29% prevalence of bacterial vaginosis (BV) among women of reproductive age [17]. In Africa the prevalence of vaginal infections is 5 million cases of chlamydia, 2 million cases of gonorrhea, and 2 million cases of syphilis among women [18]. The prevalence of BV is 50% in sub-Saharan Africa [19], 28.5% in Eastern Africa [20] and 41.9% among women in Rakai, Uganda [21]. Research shows that vaginal infections can cause symptoms like abnormal discharge, genital itching, and sores [22]. Some of these symptoms are the reason why some women would practice vaginal steaming as a hygienic practice [6].

However, there is no information comparing the presence of bacterial vaginosis between women who practice vaginal steaming and those who don’t at Mulago STI clinic. The lack of research on vaginal steaming in Uganda hinders addressing its potential implications on women’s sexual and reproductive health. The study was conducted to determine the prevalence of vaginal steaming, compare lactobacilli proportions between practitioners and non-practitioners, and identify factors associated with vaginal steaming at Mulago STI clinic.

## Methods

### Study participants and setting

This study utilized a cross-sectional design. The study was conducted for three months from November 2023 to February 2024 at Mulago STI clinic which is a national referral government facility in central Uganda located in Mulago area in Kampala capital city. The study population was women aged 18-49 years that attended the Mulago STI clinic during the data collection period who met the eligibility criteria. Women who were in their menses, those that had reached menopause and those that were one month postpartum were excluded.

### Sample size

A total of 181 participants were included in the study who met the inclusion criteria (see Fig. 1).

### Study profile

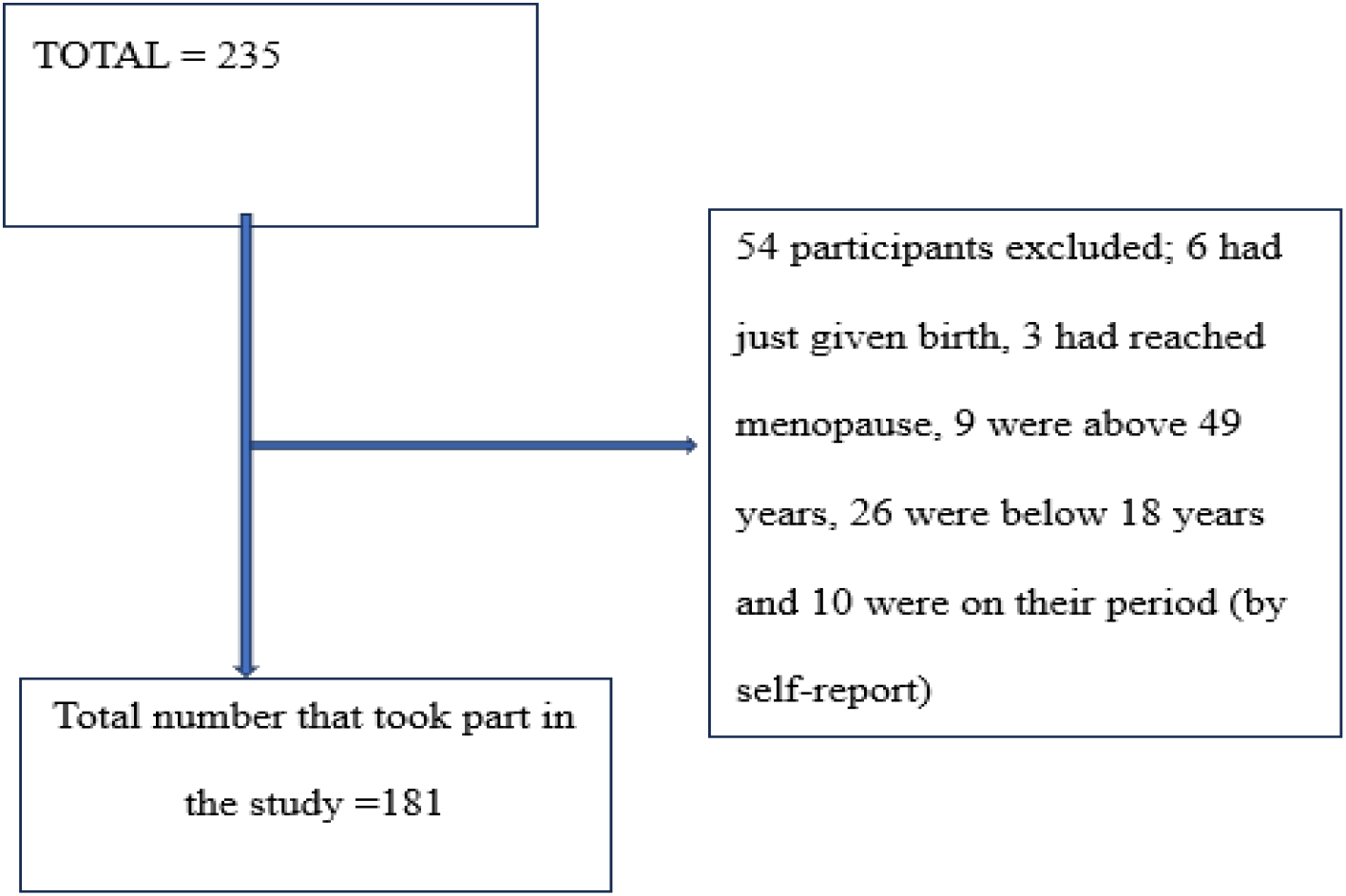
Study profile of sampled participants at Mulago STI clinic

### Sampling technique and data collection

The study was conducted from 09/092024 to 09/12/2024. Systematic sampling was applied with an interval of 2, based on an estimated clinic population of 363 and a sample size of 181. The first participant was randomly chosen from the first two eligible attendees; ineligible individuals were replaced by the next in sequence. After informed consent, data were collected using a pretested questionnaire in English or Luganda. Participants self-collected vaginal swabs after instruction, and samples were transported in cool boxes to the Infectious Disease Research Collaboration laboratory. Gram-stained slides were examined at 1000× magnification, and Nugent scoring (0–3 normal, 4–6 intermediate, ≥7 BV) classified vaginal flora.

### Study variables

Independent variables included person-related factors (age, education level, religion, occupation, knowledge of sexual and reproductive health), health system factors (access to sexual and reproductive health services), vaginal steaming status (practicing vs. not practicing), and medical history (diabetes, contraceptive use, antibiotic use within the last two weeks, urinary symptoms). For objective two, the main independent variable was vaginal steaming status, with potential confounders including medical history, person-related, and health system factors. For objective three, independent variables comprised medical history, person-related, and health system factors.

The dependent variable for objectives one and three was vaginal steaming (binary: “1” for practicing, “0” for not practicing). For objective two, the dependent variable was the presence of bacterial vaginosis (binary outcome), determined by laboratory microscopy of gram-stained vaginal swabs.

### Data analysis

Data was analysed using STATA 16.0. The descriptive statistics for categorial variables were reported as proportions and percentages whereas the continuous variables were reported as, median with their respective interquartile ranges. The prevalence of vaginal steaming was obtained as a percentage of the total number of women who practiced vaginal steaming divided by the total number of women who were enrolled into the study. Age was categorized based on the median value since it was not normally distributed and the model that had age as a categorized variable was a better model compared to that where age was a continuous variable. For the factors associated with vaginal steaming among women aged 18 to 49 years attending Mulago STI clinic, analysis was performed using a generalized linear model (modified Poisson regression model) with a log link and a Poisson distribution family using robust standard errors to cater for the model used because the prevalence was more than 10%. Data was assessed for outliers and collinearity. For the comparison of the lactobacilli proportions among women who practiced vaginal steaming and those who did not practice vaginal steaming, the lactobacilli counts were categorized into two (BV flora and normal flora). A chi square test was done to assess the crude association between vaginal steaming and bacterial vaginosis. The comparison of the lactobacilli proportions was done while adjusting for confounders using modified Poisson model with a log link and a Poisson distribution family using robust standard errors since the prevalence of bacterial vaginosis was more than 10%. The crude prevalence ratio and an adjusted prevalence ratio were calculated and compared. When the difference between the two prevalence ratios was greater than 10% then the variable was considered a confounder.

### Ethical considerations

Permission to conduct the study was obtained from 05/082024 to 05/08/2025 from the Makerere University Clinical Epidemiology Unit. Ethical clearance was obtained from the Makerere University School of Medicine Research and Ethics Committee (SOMREC). Administrative clearance was obtained from the head of Mulago National Referral Hospital. Furthermore, this study strictly adhered to ethical guidelines including the confidentiality of patients that was kept through the assignment of a barcode number on the collected sample and the data generated from the interviews was entered into a password protected laptop whose password was only known by the principal investigator of the study. In addition to the above, written informed consent was obtained from the participants before administering the questionnaire and picking any vaginal swabs from them. They were informed about their right to voluntarily be part or withdraw from the study at any one point without any penalties. Participants found to have bacterial vaginosis were informed about their condition and referred to the health care workers at Mulago STI clinic for treatment.

## Results

### DESCRIPTION OF STUDY PARTICIPANTS

#### Social demographic characteristics

The study participants had a median age of 31 years [IQR: 26, 37]. Majority of the women had attained secondary education (n = 85, 47.2%) and were employed (n = 116, 64.4%) as indicated in table 1 above.

**Table 1:** Socio-demographic characteristics of 181 study participants attending Mulago STI clinic.

| Variable | Category | Median (Q1, Q3) | Overall n (%) |
| --- | --- | --- | --- |
| Age |  | 31(26,37) |  |
| Education level | No formal education |  | 6 (3.3) |
|  | Primary |  | 30 (16.7) |
|  | Secondary |  | 85 (47.2) |
|  | Tertiary |  | 59 (32.8) |
| Occupation | Employed |  | 116 (64.4) |
|  | Unemployed |  | 64 (35.6) |

*n-number of participants, %-percentage, Q1-1st quartile, Q3- 3rd quartile*

#### Clinical characteristics of the participants

Majority of the study participants did not have diabetes (n = 170, 94.4%), 88.3% (n = 159) did not use antibiotics, 66.7 (n = 120) did not use contraceptives, and 59.1% (n =107) did not report a history of gynecological conditions. A big proportion (92.3%, n = 167) reported having had history of vaginal issues with itching being the most reported vaginal issue (41.9%, n =70) as shown in table 2 above.

**Table 2:**
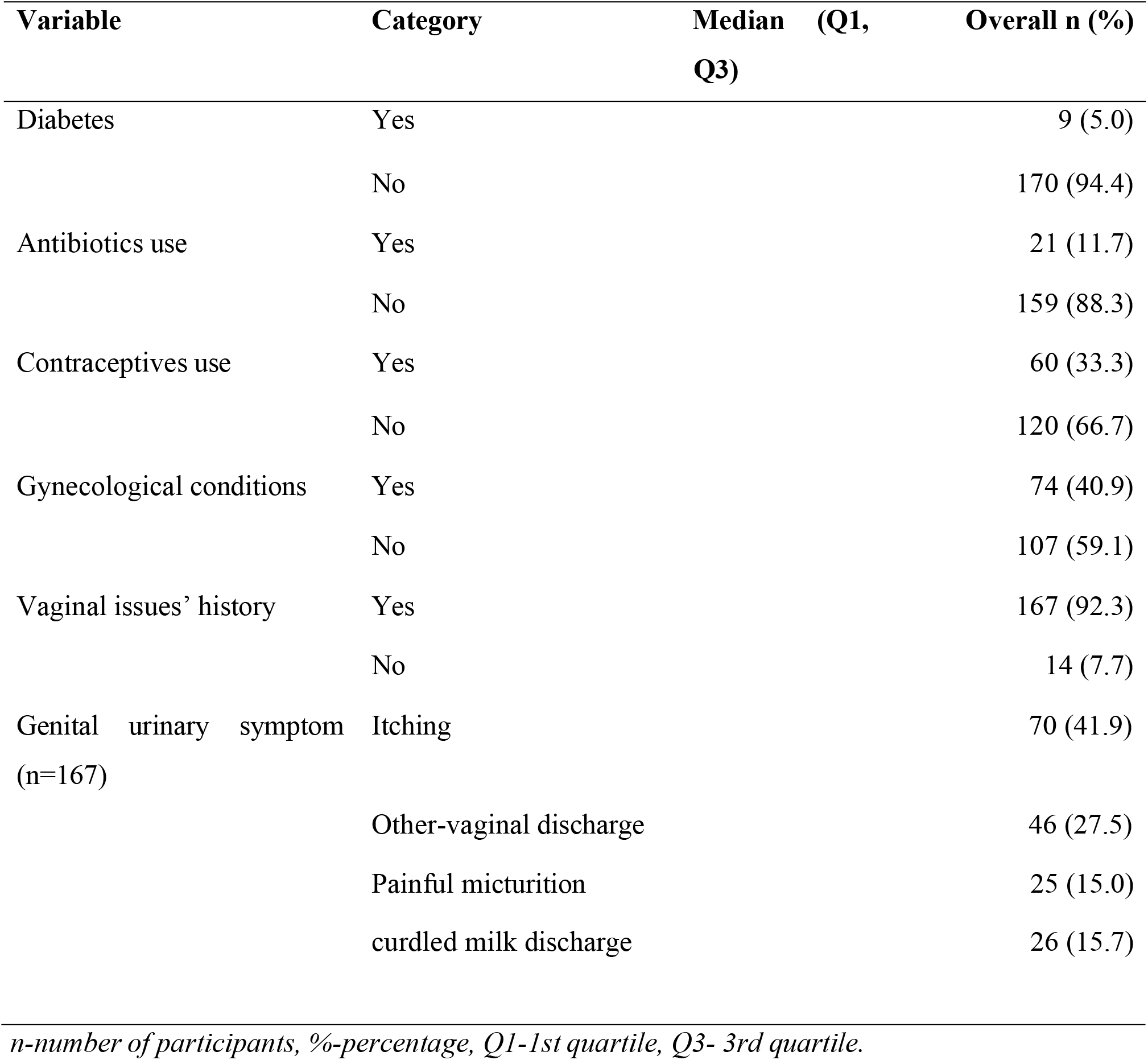
Clinical characteristics of 181 study participants attending Mulago STI clinic.

| Variable | Category | Median (Q1, Q3) | Overall n (%) |
| --- | --- | --- | --- |
| Diabetes | Yes |  | 9 (5.0) |
|  | No |  | 170 (94.4) |
| Antibiotics use | Yes |  | 21 (11.7) |
|  | No |  | 159 (88.3) |
| Contraceptives use | Yes |  | 60 (33.3) |
|  | No |  | 120 (66.7) |
| Gynecological conditions | Yes |  | 74 (40.9) |
|  | No |  | 107 (59.1) |
| Vaginal issues' history | Yes |  | 167 (92.3) |
|  | No |  | 14 (7.7) |
| Genital urinary symptom (n=167) | Itching |  | 70 (41.9) |
|  | Other-vaginal discharge |  | 46 (27.5) |
|  | Painful micturition |  | 25 (15.0) |
|  | curdled milk discharge |  | 26 (15.7) |

*n-number of participants, %-percentage, Q1-1st quartile, Q3- 3rd quartile.*

The prevalence of vaginal steaming among the 181 women attending the Mulago STI Clinic was 40.3% (95% CI: 33.4 – 47.7). There were 41.1% women who practiced vaginal steaming occasionally, 53.4% who used hot water having herbs and 78.1% who practiced vaginal steaming for medical reasons as shown in table 3 above.

**Table 3:**
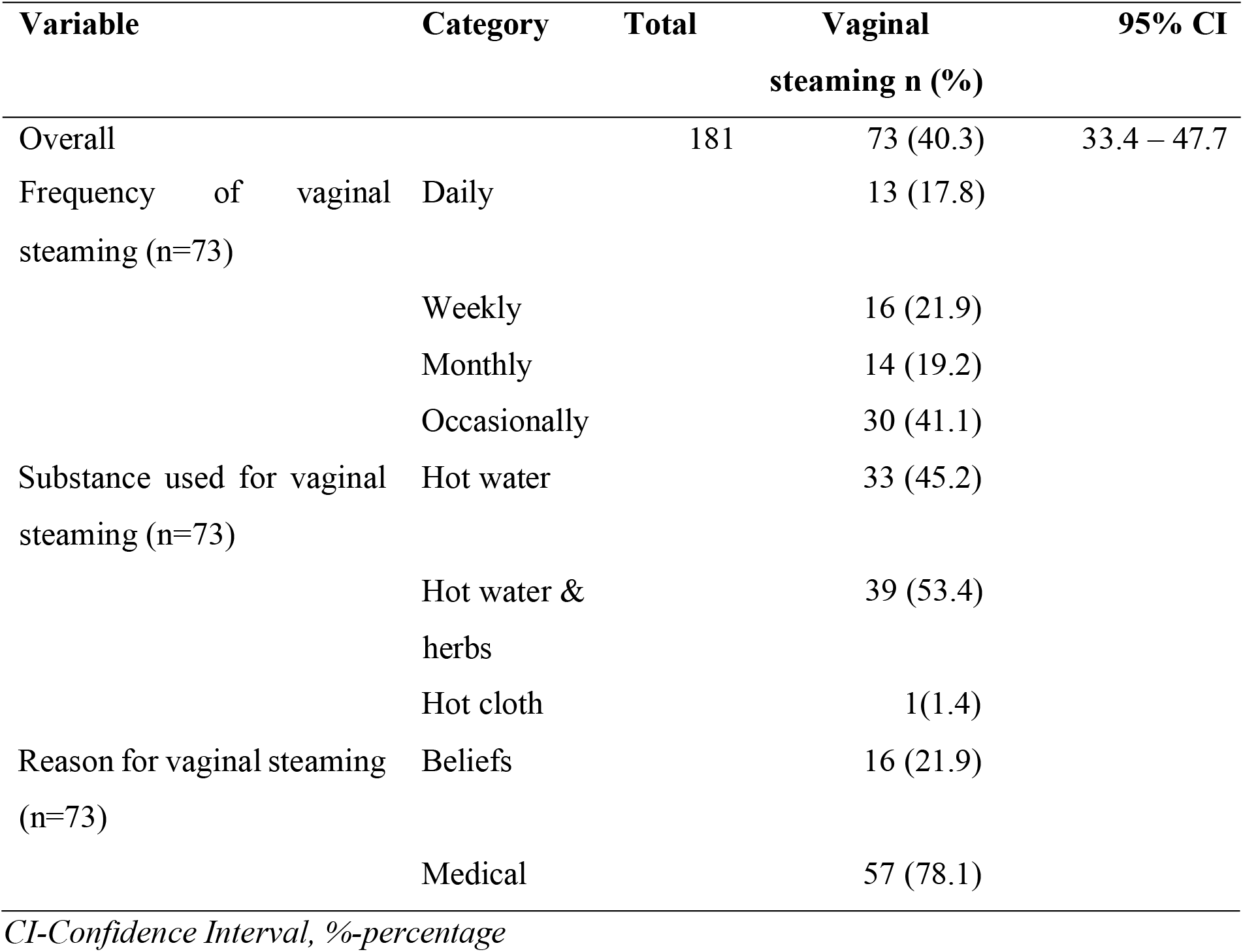
Prevalence of vaginal steaming among 181 women at Mulago STI clinic.

There was no difference in the presence of bacterial vaginosis when women who practiced vaginal steaming were compared to those who did not at Mulago STI clinic as shown in table 4 above.

**Table 4:** Comparison of the presence of bacterial vaginosis between women who practice vaginal steaming and those who did not at Mulago STI clinic.

| Bacterial vaginosis category | Practitioners of vaginal steaming N = 73, n (%) | Non-practitioners of vaginal steaming N = 108, n (%) | P-value |
| --- | --- | --- | --- |
| Presence of bacterial vaginosis | 27 (37.0) | 37 (34.3) | 0.707 |

*N is the overall number of participants in that category and n is the number of participants with bacterial vaginosis in each group, %-percentage.*

At multi variable analysis, vaginal steaming (PR-1.30, 95% CI 0.85 - 1.98) was not significantly associated with the presence of bacterial vaginosis. There was no significant interaction among the variables. However, having a history of genital urinary symptom was a confounder of the relationship between vaginal steaming and bacterial vaginosis as shown in table 5 above.

**Table 5:**
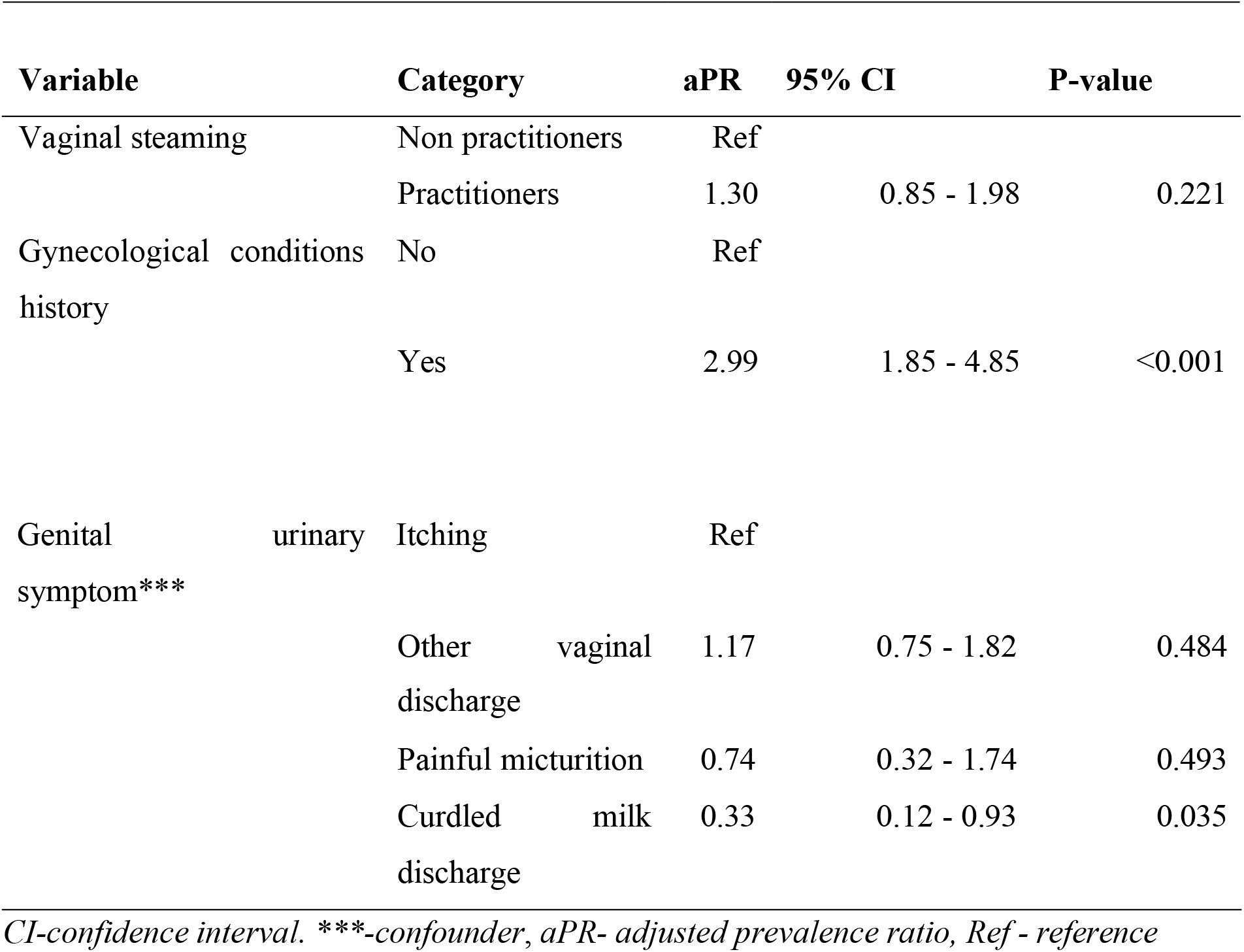
Multi variable comparison of the presence BV among women who practice vaginal steaming and those who do not at Mulago STI clinic.

#### Factors associated with vaginal steaming among 181 women at Mulago STI clinic

At multi variable analysis, history of vaginal issues (PR-0.07, 95% 0.01-0.12) and contraceptive use (PR-0.52, 95% CI 0.37 – 0.72) were statistically significantly associated with vaginal steaming as shown in Table 6 above.

**Table 6:** Multi variable analysis showing factors associated with vaginal steaming among 181 women at Mulago STI clinic.

| Variable | Category | aPR | 95% CI | P-value |
| --- | --- | --- | --- | --- |
| Contraceptives use | No | Reference |  |  |
|  | Yes | 0.52 | 0.37– 0.72 | 0.001 |
| History of vaginal issues | No | Reference |  |  |
|  | Yes | 0.07 | 0.01- 0.12 | <0.001 |

*CI-confidence interval, aPR- adjusted prevalence ratio*

## Discussion

The prevalence of vaginal steaming in this cross-sectional study was found to be 40.3% with (95% CI 33.0 – 48.0). This implies that 2 in every 5 women at Mulago STI clinic practiced vaginal steaming. This is a high prevalence since vaginal steaming practice is not recommended by any health guidelines in Uganda thus a prevalence of 0% of vaginal steaming in Uganda is the ideal. The findings are higher than the 15% reported in a house hold survey study in Mozambique [23] and this could be because the study was conducted in the community where the women in the community have less vaginal issues that may encourage vaginal steaming unlike those found at the STI clinic who may have a higher likelihood of vaginal issues which could lead them to explore alternative health practices, like vaginal steaming, as a method for managing perceived or actual health issues. Additionally, a study conducted by the Informed Health Organization found that some women cannot afford treatment for some of these infections and so resort to home remedies to treat these infections [24]. On a contrary, a multicentric cross-sectional study done in Thailand found that the prevalence of vaginal steaming was 66.9% (95% CI 62.0 - 71.5) [2] and this could be because the study was conducted in the general population where cultural practices related to vaginal steaming may be more widely accepted and routinely practiced as compared to those at the STI clinic. The study findings imply that women attending STI clinics practice vaginal steaming possibly because may believe that vaginal steaming can cleanse the vagina of any potential infections. Furthermore, some women may have limited access to health care facilities and so may resort to easily accessed practices to treat vaginal infections [17].

This prevalence report could be an underestimate or overestimate because of self-reporting as some participants could have had fears to disclose some information. However, the findings are considered true for those participants that reported having practiced vaginal steaming as they also responded to subsequent questions for details on how often they practiced vaginal steaming, reasons for the practice and the materials used.

The findings from this cross-sectional study show that 27/73 women who practiced vaginal steaming had BV whereas 37/108 women who did not practice vaginal steaming had BV. Women that practiced vaginal steaming had a 30% higher prevalence of BV than those that did not. However, the difference in the presence of BV is not significant. This implies that practicing vaginal steaming is not associated with the presence of bacterial vaginosis.

Similar findings were reported in a retrospective secondary analysis study conducted in Southwestern Uganda among women at high risk of HIV infection from towns along the trans-African highway and the shores of Lake Victoria [25]. This indifference in the presence of BV could be because, there could have been a variability in the temperature of the steam used while practicing vaginal steaming.

A prospective cohort study conducted in Japan found that higher temperatures can cause heat stress that causes a disturbance in the vaginal lactobacilli. [26]. Prospective studies conducted in Argentina found that low temperatures and normal body temperature do not affect the viability of lactobacilli respectively [27, 28].

Furthermore, the possible cause of this indifference in the presence of BV could be because vaginal steaming may not significantly alter the vaginal lactobacilli since some lactobacilli may be resilient to steam and may be affected by other factors like hormonal changes, sexual activity and hygiene practices. Similar findings were reported by a longitudinal study in the United States [29].

Having a history of gynecological issues was significantly associated with BV flora among women at Mulago STI clinic whereas having a history of genital urinary symptom was confounding vaginal steaming in the present study.

Women that had had a history of gynecological issues were 2.99 times more likely to have bacterial vaginosis when compared to those that did not have a history of gynecological issues. This implies that women with a history of gynecological issues potentially suffer from bacterial vaginosis. This association could have been due to an imbalance in the vaginal microbiota caused by the previous gynecological issue that could have caused a markable decline in beneficial Lactobacillus species and an increase in pathogenic bacteria such as *Gardnerella* and *Prevotella*, which could contribute to uterine pathology and recurrent BV. These findings are similar to those reported in a comparative study done in China [30].

Having a history of genital urinary symptoms was a potential confounder of the relationship between vaginal steaming and BV in the present study.

Women that had a history of other vaginal discharge were 1.17 times likely to have bacterial vaginosis when compared to those that had itching. This could be because women who experience frequent vaginal issues may be more likely to practice vaginal steaming as a self-treatment or traditional remedy, thereby creating a confounding effect. Similar findings were reported in a survey-based study conducted in Germany [24].

Additionally, vaginal infections often encompass a range of symptoms, including a thin white or gray vaginal discharge, odor, and pH imbalance, that are shared with BV [31]. Similar results were observed in a prospective cohort study in South Africa [32]. Furthermore, there are other vaginal infections that affect the lactobacilli counts besides bacterial vaginosis including *Neisseria Gonorrhoeae, Chlamydia Trachomatis* and *Trichomonas Vaginalis* (Martin Jr, Richardson et al. 1999, Peipert, Lapane et al. 2008, Brotman, Klebanoff et al. 2010). These findings highlight the importance of adjusting for vaginal infections and symptoms in the analysis to avoid misattributing the effects of underlying infections to vaginal steaming.

The factors that were significantly associated with vaginal steaming among women at Mulago STI clinic were having experienced vaginal issues and contraceptive use.

Women that used contraceptives had a 48% lower prevalence of vaginal steaming than those that did not. This could be because some women practice vaginal steaming for perceived fertility-related benefits and contraceptive users may not feel the need for these benefits, especially if they’re not trying to conceive. Similar findings were reported in a multi-site systematic review done in Australia, Canada, New Zealand, the UK, the US [5]. However, the results from the present study varied from those by a survey conducted in France which suggested that some women who use contraceptives may practice vaginal steaming [33]. This could be because the present study was conducted in a hospital with a controlled environment with fewer external distractions, allowing for standardized administration of questionnaires to the participants.

Women that had experienced vaginal issues had a 93% lower prevalence of vaginal steaming than those that had not experienced vaginal issues. This could be because some women who may have experienced vaginal issues in their past, such as infections, irritation, or discomfort, may prefer evidence-based medical treatments and avoid unproven alternative practices like vaginal steaming. Similar findings were reported in a survey conducted in Netherlands [34]. However, the findings of the present study are different from those reported by cross sectional study in China [35] could be because the later study was conducted among female sexual workers who may opt for various co-medications to relieve some vaginal infections as fast as possible to keep doing their work.

### Limitations of the study

Data were collected using an interviewer-administered questionnaire, which may have introduced social desirability and reporting biases for variables such as vaginal steaming and diabetes status, potentially leading to under- or overestimation of their effects. Self-report bias was mitigated through rapport building, accurate translation, and cross-checking of responses. Selection bias was possible as postmenopausal women more prone to bacterial vaginosis were excluded, although they comprised only 1.5% of the clinic population, limiting its impact.

The cross-sectional design precluded establishing temporal relationships between bacterial vaginosis, vaginal steaming, and covariates, and not all potential confounders (e.g., diabetes mellitus, hygiene practices) were assessed. Findings may not be generalizable beyond Mulago STI Clinic. Nevertheless, these limitations are unlikely to have substantially influenced the results, which are considered valid.

## Conclusion

Vaginal steaming is a common practice among women attending Mulago STI clinic, with nearly 2 in every 5 women reporting engagement in the practice. There was no difference in the presence of bacterial vaginosis among women who practiced vaginal steaming when compared to those who did not practice vaginal steaming. Having a history of vaginal issues and contraceptive use were significantly associated with practicing vaginal steaming among women at Mulago STI clinic

### Implication for research and practice

Patients should be informed that, despite the lack of association between vaginal steaming and bacterial vaginosis, the practice is not recommended in any health guidelines, and only Ministry of Health–approved medicines should be used for treatment. Healthcare workers in STI and family planning clinics should screen for vaginal steaming and advise against its use for managing vaginal infections. The Ministry of Health should implement targeted screening and treatment for bacterial vaginosis, develop counselling guidelines for healthcare providers, and support further longitudinal and qualitative research to explore the motivations, safety, and long-term outcomes of vaginal steaming.

## Data Availability

All data produced in the present study are available upon reasonable request to the authors.

https://doi.org/10.5281/zenodo.21840522

## List of abbreviations

BV: Bacterial Vaginosis.
HIV: Human Immunodeficiency Virus.
MOH: Ministry of Health.
OR: odds ratio.
SOM-REC: School of Medicine Research and Ethics Committee.
SSA: Sub–Saharan Africa.
STDs: Sexually Transmitted Diseases.
STI: Sexually transmitted infections.
UK: United Kingdom.
USA: United States of America.
Vlv: volume by volume.
WHO: World Health Organization.

## Declarations Acknowledgements

We appreciate the support rendered by Infectious Disease Research Collaboration for the laboratory, Mulago National referral hospital for the staff and study site. We thank all the participants in this study, and Makerere University College of Health Sciences Clinical Epidemiology Unit for the opportunity to contribute to the knowledge in this field.

## Author information

Makerere University College of Health Sciences Clinical Epidemiology, Kampala, Uganda. Dorothy Kakai, Racheal Namutale, Twinamasiko Nelson, Mugerwa Jovan, Enos Kigozi, Akinyi Lilian, Muyunga Anthony, Bagaya Jamidah, Mutesi Bushira.

Makerere University College of Health Sciences Obstetrics and Gynecology department, Kampala, Uganda.

Sarah Nakubulwa.

Makerere University College of Health Sciences Microbiology department, Kampala, Uganda.

Kajumbula Henry.

## Contributions

Conceptualization: DK, KH, SN

Data curation: DK

Formal analysis: MB, DK, EK, TN, AL

Funding acquisition: DK

Investigation: DK, KH

Methodology: DK, MB, BJ, RN

Validation: DK, EK, RN

Supervision: HK, SN

Project administration: DK, RN

Writing original draft: DK, KH, SN

Writing review & editing: TN, RN, KE, AL, BJ, MB, KH, SN

## Corresponding author

Correspondence to Dorothy Kakai

## Ethics declarations

Ethics approval and consent to participate

Written informed consent was obtained from participants to participate in the study. Study procedures were approved by the Makerere University School of Medicine Research and Ethics Committee and Mulago National Referral Hospital Research Ethics Committee. Data were collected in accordance with international conventions and guidelines on research involving human subjects such as the declaration of Helsinki.

## Availability of data and materials

The datasets used and analyzed during the current study are available from the corresponding author on reasonable request.

## Consent for publication

Not applicable.

## Rights and permissions

**Open Access** This article is licensed under a Creative Commons Attribution-Non-Commercial-No Derivatives 4.0 International License, which permits any non-commercial use, sharing, distribution and reproduction in any medium or format, as long as you give appropriate credit to the original author(s) and the source, provide a link to the creative.

